# Untargeted Metabolome Atlas for Sleep Phenotypes in the Hispanic Community Health Study/Study of Latinos

**DOI:** 10.1101/2024.05.17.24307286

**Authors:** Ying Zhang, Brian W Spitzer, Yu Zhang, Danielle A Wallace, Bing Yu, Qibin Qi, Maria Argos, M Larissa Avilés-Santa, Eric Boerwinkle, Martha L Daviglus, Robert Kaplan, Jianwen Cai, Susan Redline, Tamar Sofer

## Abstract

Sleep is essential to maintaining health and wellbeing of individuals, influencing a variety of outcomes from mental health to cardiometabolic disease. This study aims to assess the relationships between various sleep phenotypes and blood metabolites. Utilizing data from the Hispanic Community Health Study/Study of Latinos, we performed association analyses between 40 sleep phenotypes, grouped in several domains (i.e., sleep disordered breathing (SDB), sleep duration, timing, insomnia symptoms, and heart rate during sleep), and 768 metabolites measured via untargeted metabolomics profiling. Network analysis was employed to visualize and interpret the associations between sleep phenotypes and metabolites. The patterns of statistically significant associations between sleep phenotypes and metabolites differed by superpathways, and highlighted subpathways of interest for future studies. For example, some xenobiotic metabolites were associated with sleep duration and heart rate phenotypes (e.g. 1H-indole-7-acetic acid, 4-allylphenol sulfate), while ketone bodies and fatty acid metabolism metabolites were associated with sleep timing measures (e.g. 3-hydroxybutyrate (BHBA), 3-hydroxyhexanoylcarnitine (1)). Heart rate phenotypes had the overall largest number of detected metabolite associations. Many of these associations were shared with both SDB and with sleep timing phenotypes, while SDB phenotypes shared relatively few metabolite associations with sleep duration measures. A number of metabolites were associated with multiple sleep phenotypes, from a few domains. The amino acids vanillylmandelate (VMA) and 1-carboxyethylisoleucine were associated with the greatest number of sleep phenotypes, from all domains other than insomnia. This atlas of sleep-metabolite associations will facilitate hypothesis generation and further study of the metabolic underpinnings of sleep health.

## Introduction

Sleep plays an important role in the health and wellbeing of individuals. Insufficient quality, timing, and duration of sleep have a major public health impact, and are associated with daytime sleepiness, poor mental health, impaired cognitive function, and increased risk of cardiovascular morbidity and mortality (1–3). Sleep is increasingly recognized as a crucial factor in cardiovascular health, evident by the addition of sleep duration to the “life’s essential 8” metric developed by the American Heart Association(4). In addition to sleep duration, measures of suboptimal sleep, such sleep disturbances and quality (or insomnia symptoms), irregularity of sleep timing, sleep fragmentation, and sleep disordered breathing (SDB), are also associated with poor health outcomes (5). In fact, there is growing recognition of the importance in measuring and characterizing multi-dimensional sleep health— a framework that concurrently considers these varied aspects of sleep (6–8).

Despite the strong epidemiological evidence observed in many cohort and clinical studies for the connection between suboptimal sleep health and increased risks for poor health outcomes, the biology and physiology behind these links are not fully understood. While many sleep behaviors and outcomes share some underlying genetic and physiological pathways (9–11), or have, potentially bidirectional, causal relationships (12), there may also be distinct mechanisms that underlie specific sleep disturbances or sleep subtypes (13–15). Untangling these shared and distinct mechanisms underlying sleep phenotypes has the potential to inform sleep health intervention efforts.

Biological sampling to measure molecular markers of health, such as in metabolomics, can be used to investigate the mechanisms and pathways underpinning sleep phenotypes. Metabolites are small molecules produced in the formation and/or breakdown of endogenous or exogenous substances and are oriented at the closest layer to phenotypes compared to other underlying biochemical layers (e.g., genome, transcriptome and proteome). The increasing availability of large datasets with untargeted metabolomics profiling has unveiled metabolic outcomes and correlates of numerous health phenotypes, including sleep measures (16–31). A large-scale study of metabolites in relation to sleep phenotypes may shed light on how underlying biological processes may converge and differ among common sleep phenotypes, the complex interplay between sleep and the metabolic environment, and, ultimately, potential interactions among sleep disorders and progression of cardiometabolic and other health conditions. Here, we study the associations between a range of sleep phenotypes and the metabolic environment in a large population-based study using a high-dimensional set of measured metabolites. We create an “atlas” – a resource for the sleep research community that will facilitate hypotheses formulation and accelerate studies on sleep and its association with other health outcomes.

Our study has taken a comprehensive approach, covering key sleep phenotypes including sleep disordered breathing (SDB), sleep duration, sleep timing, insomnia symptoms, and heart rate (HR) during sleep. Each category of sleep phenotypes provides a different perspective on sleep, while together may highlight some shared biological processes within this complex physiological phenomenon. We also conducted network analysis to better understand the interconnectedness between multiple sleep phenotypes and metabolites –representing significant associations as links in a bipartite network allows for simultaneous visualization of many associations, enabling researchers to perceive connectivity patterns that might otherwise be obscured when looking at the individual relationships. By reporting a large number of associations between metabolites and sleep phenotypes, this resource may provide researchers with a starting point for more targeted inquiries into the metabolic environment changes induced by sleep disorders, and facilitate hypothesis generation for future metabolomic sleep research. This may ultimately contribute to our understanding of the pathogenesis of sleep disorders and pave the way for developing more effective diagnostic and therapeutic strategies.

## Methods

### The Hispanic Community Health Study/Study of Latinos

The Hispanic Community Health Study / Study of Latinos (HCHS/SOL) is a prospective community-based cohort study of 16,415 Hispanic/Latino individuals aged 18–74 years at the baseline examination (2008-2011). Study participants were selected using a multi-stage stratified random sampling from four geographic regions: Bronx NY, Chicago IL, Miami FL, and San Diego CA (32,33). Description of major ancillary studies and findings in the context of cardiovascular health is provided in a prior publication (34). Fasting blood samples were collected at the baseline examination, and within the subsequent week, 14,440 of these participants underwent an evaluation for SDB using a validated Type 3 home sleep apnea test (ARES Unicorder 5.2; B-Alert, Carlsbad, CA) that measured nasal airflow, position, snoring, heart rate and oxyhemoglobin saturation with measures of SDB scored by a central reading center as detailed previously (35). All sleep phenotypes used and their definitions are provided in Supplementary Note 1.

### Metabolomics profiling

From those who attended the HCHS/SOL baseline assessment and also underwent genotyping (12,803 of the study individuals (36)), 4,002 individuals were selected at random for metabolomics profiling of fasting serum samples collected at baseline (metabolomics batch 1, processed in 2017). In 2021, additional 2,368 serum samples from 2,330 participants, also collected at baseline, were profiled in a second metabolomics batch 2. Serum samples were stored at -70°C at the HCHS/SOL Core Laboratory at the University of Minnesota until analysis by Metabolon, Inc. (Durham, NC) in 2017 (batch 1) and 2021 (batch 2). Serum samples were then extracted and prepared using Metabolon’s standard solvent extraction method. Extracts were split into a five fractions to use in four liquid chromatography-mass spectrometry (LC-MS)-based metabolomic quantification platforms (two reverse phase methods with positive ion mode electrospray ionization (EI), one reverse phase method with negative ion mode EI, and one hydrophilic interaction liquid chromatography with negative ion mode EI), with the fifth fraction reserved for backup. Instrument variability was assessed by calculating the median relative standard deviation (SD) for the internal standards added to each sample prior to injection into the mass spectrometers. Overall process variability was determined by calculating the median relative SD for all endogenous metabolites (i.e., non-instrument standards) present in 100% of the technical replicate samples.

### Metabolomic data pre-processing

Preprocessing of the metabolomic data is described in Supplementary Figure S1. First, we removed batch 2 individuals who overlapped with batch 1 and replicate samples from the same individuals, resulting in 2,178 remaining batch 2 observations. We then computed percentages of missing values of each metabolite in each batch separately. We excluded metabolites with missing values in more than 75% of the individuals in either batch. For xenobiotic metabolites (metabolite annotation was provide by Metabolon), we assumed that missing values were due to concentrations below the minimum detection limits, thus imputed the missing values for each metabolite with half of the lowest non-missing value of that metabolite across the sample within the batch. For non-xenobiotic metabolites, we applied multiple imputation using the futuremice function from the R mice package (version 3.15.0) that implements fully conditional imputation in a computationally efficient manner (using parallelization). Each variable in the imputation dataset is imputed (if it has missing values) using a model that predicts potential values based on other variables in the dataset. The dataset was imputed 5 times to generate 5 completed datasets. Differences between the computed datasets are due to randomness (e.g. random residuals added to the predicted values of a variable). We imputed metabolites, separately in each batch, together with a set of covariates that are strongly linked to the metabolic environment: age, sex, BMI, waist-to-hip ratio, fasting insulin, fasting glucose, type 2 diabetes status (American Diabetes Association definition: fasting glucose >=126 mg/dL, or post-OGTT glucose >=200 mg/dL or A1C>=6.5%, or self-report of diabetes), estimated glomerular filtration rate, and lipid measures – total cholesterol, triglycerides, high- and low-density lipoprotein measures. We then rank normalized the 768 metabolite measures passing quality control in each batch separately and each imputed dataset separately, and finally, aggregated data from the two batches, such that the “first” imputed batch 1 dataset was aggregated with the “first” imputed batch 2 dataset, and so on. Supplementary Note 2 describes the analysis we performed to inform the metabolite imputation strategy.

### Sleep phenotypes and modeling approach

We used 40 sleep phenotypes from 5 domains, described in detail in Supplementary Note 1. In brief, these included: self-reported sleep duration (sleep duration during weekdays and weekend days, average sleep duration during, short and long sleep during weekdays), heart rate during sleep (minimum, maximum, average, and standard deviation of HR), insomnia (the women health initiative insomnia rating scale (WHIIRS), its component questions, sleeping pill use, and daytime and excessive daytime sleepiness, measured via the Epworth sleepiness scale), sleep disordered breathing (respiratory event indices, measures of oxyhemoglobin saturation during sleep, measures of respiratory event lengths, as well as self-reported snoring), and sleep timing (bed time and wake time as well as sleep midpoint during weekend and weekend days, and social jetlag – the difference in sleep midpoint between weekend and weekdays). In figures and tables, these domains are referred to as Duration, HR, Insomnia, SDB, and Timing. Phenotypes in the sleep timing category were analyzed as “circular” variables, i.e., to avoid bias due to day thresholding (i.e., defining a day by midnight) and to account for the fact that 11:59PM (23:59) is adjacent to 12:01AM (00:01). Other sleep phenotypes were treated as linear or binary (when dichotomized). Two nested conceptual models were used in the analyses. Model 1 adjusted for batch number, demographic, and baseline clinical variables, including age, sex, field center, Hispanic/Latino background (Mexican, Puerto Rican, Cuban, Central American, Dominican, South American, and other/multi) and body mass index (BMI). Model 2 further adjusted for lifestyle variables – alcohol use (never, former, current), cigarette use (never, former, current), total physical activity (MET-min/day, computed based on self-reported time spent doing physical activities), and diet (Alternative Healthy Eating Index 2010, computed based on 24 hours dietary recall data) in addition to Model 1 covariates. Sleep phenotypes were used as exposures with metabolites as outcomes. When testing the association of a circular phenotype (i.e., sleep timing) with a metabolite, we first converted the sleep timing phenotype into radians (with 12am serving as the 0) and then computed the sine and cosine of these radians. We used the sine and cosine terms as predictors in the regression (37), and tested their association with the metabolite using the multivariate Wald test, accounting for two predictors and their estimated covariance. All association analyses were performed using the *survey* R package (version 4.1) to account for the HCHS/SOL sampling design and provide effect estimates relevant for the HCHS/SOL target population.

### Estimating associations between metabolites and sleep phenotypes

We separately assessed the association between each metabolite’s concentration level, as an outcome, and each sleep phenotype, as a predictor, using Model 1 and Model 2 covariates as described above, in a single metabolite association analysis. Xenobiotic metabolites were imputed once, while missing values in other metabolites values were imputed 5 times. Thus, we estimated the associations of non-xenobiotic metabolites with sleep phenotypes 5 times (using each of the completed datasets), and then combined the resulting estimated associations. Here we treated the linearly-modeled sleep phenotypes differently from the circular ones. The estimated metabolite associations with linearly-modeled sleep phenotypes were combined using Rubin’s rule (38). Sleep timing phenotypes cannot be combined in the same manner, because there is no method to combine the covariance between the sine and cosine terms across several completed datasets. Instead, we focused on testing and aggregated the p-value of the multivariate Wald test using the aggregated Cauchy association test (ACAT; (39)). The ACAT test was developed in the context of genetic association analyses, but it is appropriate for our settings, because it allows for the aggregated tests to be based on correlated data. Finally, after combining association results so we had one p-value per sleep phenotype-metabolite association (per model), we implemented the Benjamini-Hochberg method to control the false discovery rate (FDR) for multiple testing across all metabolites in all models for each sleep phenotype (40). Any association that resulted in an FDR-adjusted p-value<0.05 in Model 1 was considered statistically significant. In secondary analysis, we also performed sex-stratified analyses using the same analytic approach. For descriptive purposes, we computed the number of sleep traits that each metabolite was associated with at the FDR<0.05 Model 1 threshold, and identified the top 10% metabolites based on number of sleep trait associations.

We summarized the associations between sleep phenotypes, individually and categorized by domains, and metabolites, individually and categorized by pathways. Similarities of these associations between a pair of phenotypes or domains were estimated with the Dice Similarity Coefficient (DSC) (41) defined as

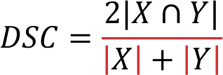

Where *X* and *Y* represent the set of metabolites with statistically significant associations (FDR<0.05) with two sleep phenotypes or domains. DSC takes values between 0 and 1 where 0 indicates no similarity while 1 indicates full overlap between the two sets.

### Bipartite network analysis

Bipartite network is a type of network in which there are two groups of nodes (here: sleep phenotypes and metabolites), and links, or edges, can only exist between the two types of nodes but not within a group of nodes. We constructed a bipartite network using the sleep phenotypes and metabolites, where an edge was added between a sleep phenotype-metabolite pair if their association was statistically significant (FDR-adjusted *p*-value<0.05) in Model 1 association analysis. The network is built based on an incidence matrix, with rows corresponding to sleep phenotypes, columns represent metabolites, and the *i*, *j* cell in this matrix has a value of 1 if the *i*th sleep phenotype has a statistically significant association with the *j*th metabolite, and 0 otherwise. We then aggregated metabolites by sub- and super-pathway, and sleep phenotypes by domain, which results in a consolidated incidence matrix in which a value of 1 in cell(*m,n)* indicating any statistically significant association between the *m*th sleep phenotype domain and the *n*th metabolite sub- and super-pathway and a weight matrix documenting the number of total statistically significant associations between the *m*th sleep phenotype domain and the *n*th metabolite sub- and super-pathway. Several network property metrics were computed to offer insights into the structure and associations of the sleep domain – metabolite sub- and super-pathway network.

For visualization, we converted the bipartite network into a univariate network and visualized the network using the Fruchterman-Reingold force-directed algorithm, which optimizes the placement of nodes based on connectivity similarity between nodes, where similar connectivity is reflected as proximity of nodes (42), providing an intuitive spatial representation of the network structure.

### Statistical software

All analyses were done in R version 4.2.3. The function svyglm from the survey package was used for survey-weighted generalized linear regression models. The car package (version 3.1) was used for multivariate Wald test. The bipartite package (version 2.18) was used for bipartite network analysis, and igraph (version 1.5) and ggnetwork (version 0.5) packages were used for visualizing network graph.

## Results

### Study sample characteristics

Table 1 characterizes the HCHS/SOL target population (using means and percentages weighted to account for study participation), with more comprehensive data provided in Supplementary Table S1. The analytic sample, batch 1 and batch 2 metabolomic dataset combined, included 6,180 participants with a mean age of 44.31 years (SD=15.27), of whom 40.1% were males, 20.7% were non-alcohol users, 58.5% never smoked, and 11.7% had moderate-to-severe OSA (REI3>=15). The average sleep duration was 7.98 hours (SD=1.46), while 22.7% reported restless or very restless sleep on a typical night in the last month, and 15.8% reported excessive sleepiness (ESS>=10). Batch 2 participants were older compared to batch 1 – the mean age is 41.54 years (SD) in batch 1 and 51.11 years (SD) in batch 2. The baseline rates of diabetes and hypertension were higher in batch 2 (diabetes: 29.4%; hypertensions: 44.1%) compared to batch 1 (diabetes: 20.1%; hypertensions: 31.5%), consistent with the age difference between the two batches.

**Table 1:** HCHS/SOL target population characteristics by sex, batch, and overall.

|  |  |  |  |  |  |
| --- | --- | --- | --- | --- | --- |
|  | Female | Male | Batch 1 | Batch 2 | Overall |
| **n** | 3704 | 2476 | 4002 | 2178 | 6180 |
| **Age (mean (SD))** | 44.98<br>(15.17) | 43.56 (15.35) | 41.54<br>(15.18) | 51.11<br>(13.21) | 44.31<br>(15.27) |
| **Gender = Male (%)** | 0 ( 0.0) | 2476 (100.0) | 1705<br>(42.6) | 771<br>(35.4) | 2476<br>(40.1) |
| **Hispanic/Latino background (%)** |  |  |  |  |  |
| **BMI (mean (SD))** | 30.15<br>(6.69) | 28.78 (5.37) | 29.42<br>(6.27) | 29.72<br>(5.80) | 29.50<br>(6.14) |
| **Current alcohol drinking (%)** | 1401<br>(37.9) | 1490 ( 60.2) | 1973<br>(49.3) | 918<br>(42.3) | 2891<br>(46.8) |
| **Current smoking (%)** | 596<br>(16.1) | 668 ( 27.1) | 863<br>(21.6) | 401<br>(18.5) | 1264<br>(20.5) |
| **Physical activity (MET-min/day) (mean (SD))** | 424.88<br>(714.71) | 879.70<br>(1174.88) | 697.66<br>(1020.41) | 496.61<br>(880.66) | 639.60<br>(986.25) |
| **The Alternate Healthy Eating Index (2010) (mean (SD))** | 46.79<br>(7.42) | 48.74 (7.47) | 47.28<br>(7.52) | 48.75<br>(7.36) | 47.71<br>(7.50) |
| **OSA status = OSA (%)** | 260 ( 7.9) | 383 ( 17.4) | 388<br>(10.9) | 255<br>(13.0) | 643<br>(11.7) |
| **Baseline Diabetes status (ADA) = Yes (%)** | 863<br>(23.3) | 579 ( 23.4) | 803<br>(20.1) | 639<br>(29.4) | 1442<br>(23.3) |
| **Baseline Hypertension status = Yes (%)** | 1357<br>(36.6) | 865 ( 34.9) | 1261<br>(31.5) | 961<br>(44.1) | 2222<br>(36.0) |
| **Sleep Duration: Work/School Days (in hours) (mean (SD))** | 7.89<br>(1.66) | 7.75 (1.56) | 7.82<br>(1.62) | 7.83<br>(1.60) | 7.82<br>(1.62) |
| **Womens's Health Initiative Insomnia Rating Scale (WHIIRS) total score (mean (SD))** | 7.64<br>(5.58) | 6.16 (5.13) | 6.78<br>(5.37) | 7.34<br>(5.54) | 6.95<br>(5.42) |
| **Epworth Sleepiness Scale (ESS) total score (mean (SD))** | 5.44<br>(4.71) | 5.87 (4.87) | 5.62<br>(4.71) | 5.70<br>(4.99) | 5.64<br>(4.79) |
| OSA was defined as respiratory event index >=15. |  |  |  |  |  |
| Means and percentages have been weighted to provide values representative of the HCHS/SOL target population. |  |  |  |  |  |

### Results from sleep phenotype-metabolite association analysis

The single metabolite association analysis was conducted in pair-wise fashion between 40 sleep phenotypes from five domains (i.e., sleep duration, HR during sleep, insomnia, SDB, and sleep timing) and 768 metabolites, including 113 unknown metabolites and 77 xenobiotic metabolites. Of the phenotypes, 35 sleep phenotypes had statistically significant associations with at least one metabolite (Table 2). When limited to Model 1, the median number of significant associations for each sleep phenotype is 16.5 (range: 0 - 304), corresponding to 2.15% (range: 0% – 39.58%) of all tested metabolites. The number of statistically significant associations is much lower among dichotomized sleep phenotypes (median: 5; range: 0 – 73) compared to non-dichotomized sleep phenotypes (median: 37; range: 0 – 304), likely corresponding to loss of power due to dichotomization. Phenotypes from the heart rate during sleep domain had the highest number of statistically significant associations with metabolites (median: 174, range: 2 - 304), followed by the sleep timing domain (median: 117, range: 64 - 197). The SDB domain had the lowest number of statistically significant associations with tested metabolites (median: 8, range: 0 – 68) (Supplementary Table S2). Figure 2 visualizes the strength of associations (i.e., negative logarithm of the FDR-adjusted *p*) between sleep phenotypes and individual metabolites grouped by superpathway. Compared to the sex-stratified analysis, sex-combined analysis tends to identify more statistically significant associations between sleep phenotypes and metabolites (Figure 3), in accordance with the higher power due to larger sample size of the sex-combined analysis.

**Figure 1.**
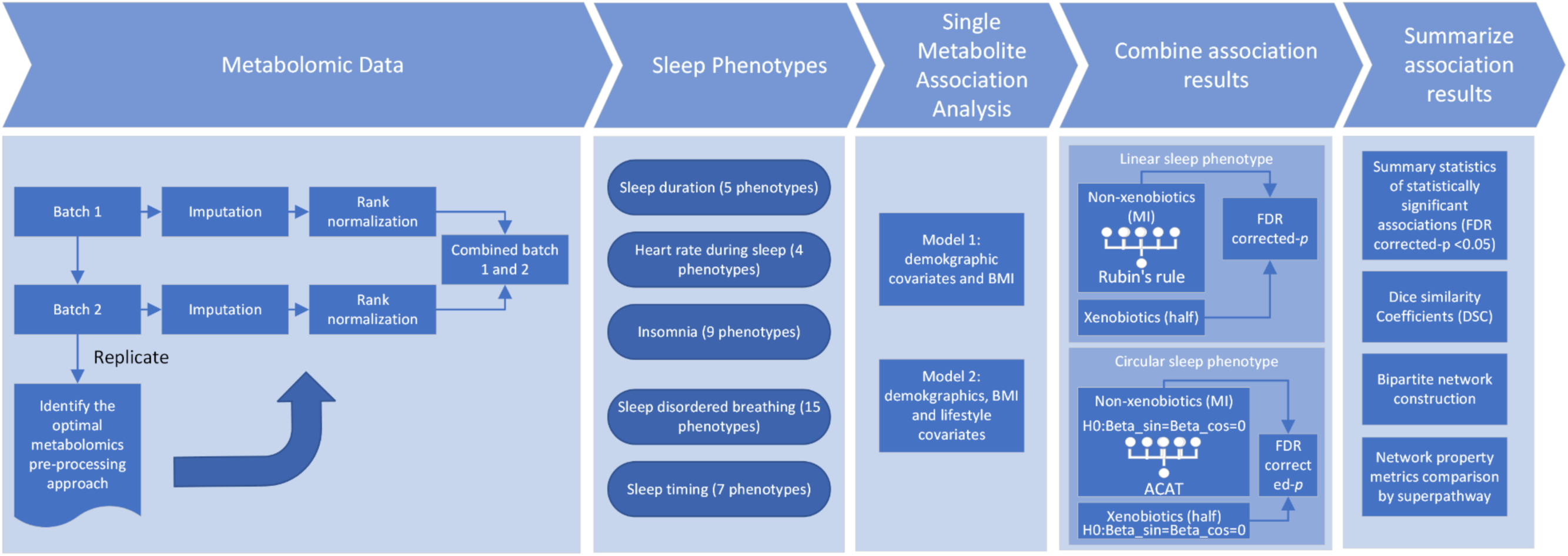
Study design diagram.

**Figure 2.**
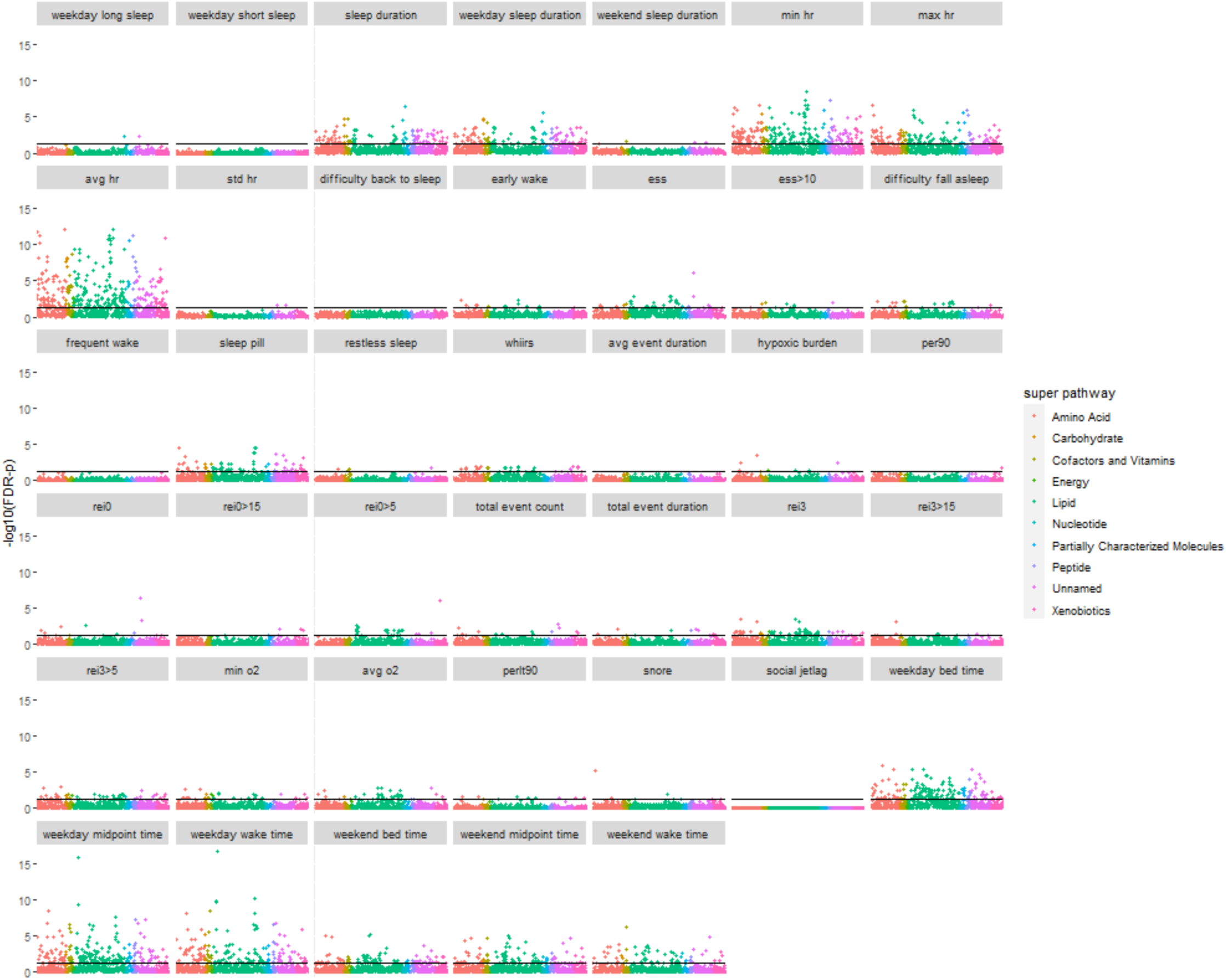
Scatter plot of single metabolite association analysis with sleep phenotypes. -log10(FDR-*p*) is based on FDR-adjusted *p* in which the raw *p* was derived by accounting for the complex sampling design-based degrees of freedom, using adjusted standard errors to compute the t-statistic in single metabolite association analysis with each sleep phenotype as dependent variables, while the FDR adjustment was based on the Benjamini-Hochberg method to control false discovery rate (FDR) for multiple testing among all metabolites in all models for each sleep phenotype. Black horizontal line indicates FDR-p=0.05.

**Figure 3.**
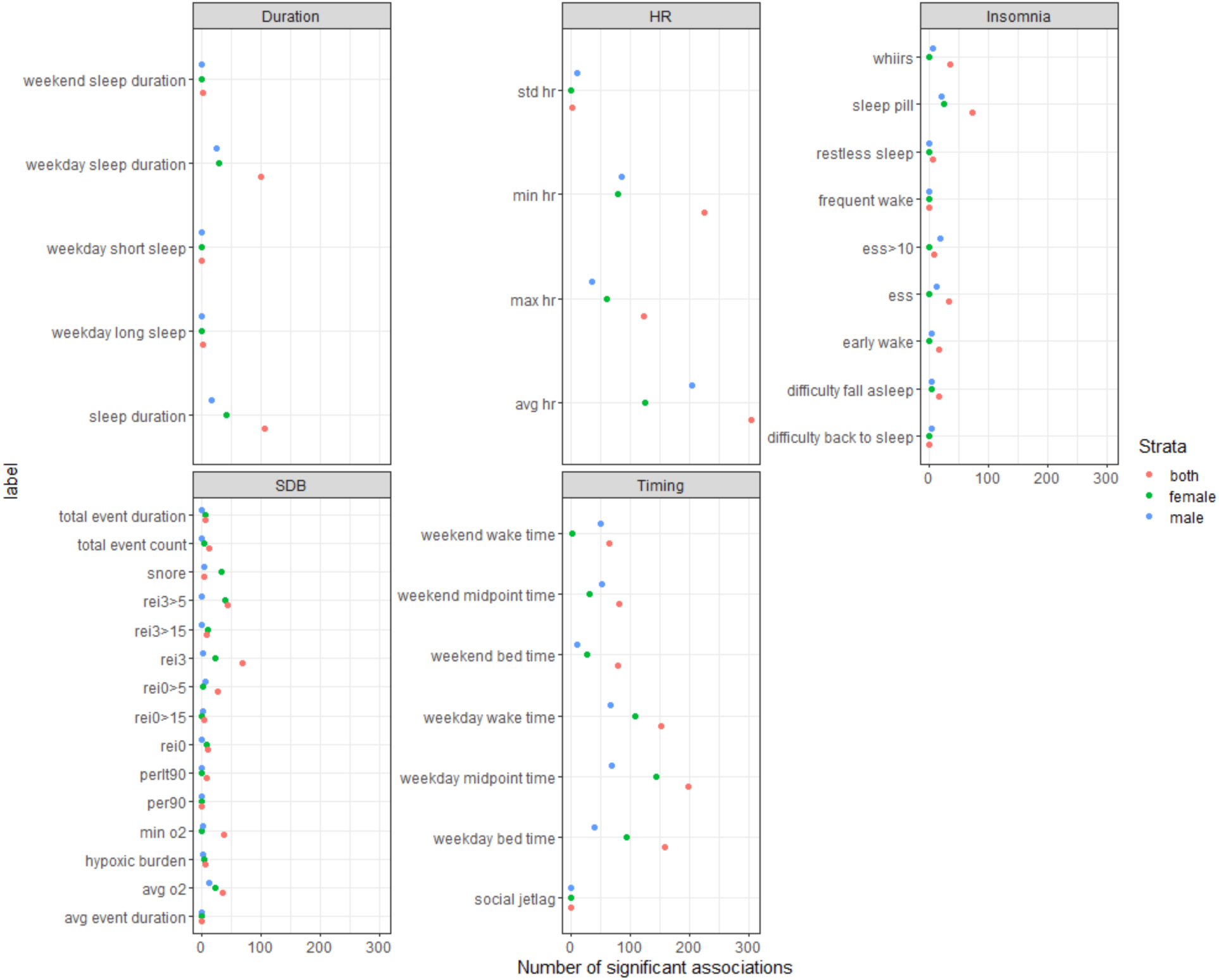
Number of statistically significant associations between metabolites and sleep phenotypes by domain and sex strata. Associations between metabolite and sleep phenotype are identified as statistically significant based on FDR-p<0.05. The raw *p*-value was derived by accounting for the complex sampling design-based degrees of freedom, using adjusted standard errors to compute the t-statistic in single metabolite association analysis with each sleep phenotype as dependent variables. The FDR adjustment was based on the Benjamini-Hochberg method to control false discovery rate (FDR) for multiple testing among all metabolites in all models for each sleep phenotype.

**Table 2:** Statistically significant metabolite associations by sleep phenotype.

| **Sleep phenotype** | **Number of statistically significant associations** | **Percentage of statistically significant associations (%)** | **Sleep phenotype domain** | **Sleep phenotype dichotomized or continuous** |
| --- | --- | --- | --- | --- |
| weekday long sleep | 2 | 0.26 | Duration | Dichotomized |
| weekday short sleep | 0 | 0.00 | Duration | Dichotomized |
| sleep duration | 107 | 13.93 | Duration | Continuous |
| weekday sleep duration | 101 | 13.15 | Duration | Continuous |
| social jetlag | 0 | 0.00 | Duration | Continuous |
| weekend sleep Duration | 3 | 0.39 | Duration | Continuous |
| min HR during sleep | 225 | 29.30 | Heart rate (HR) | Continuous |
| max HR during sleep | 123 | 16.02 | Heart rate (HR) | Continuous |
| avg HR during sleep | 304 | 39.58 | Heart rate (HR) | Continuous |
| std HR during sleep | 2 | 0.26 | Heart rate (HR) | Continuous |
| difficulty back to sleep | 0 | 0.00 | Insomnia | Dichotomized |
| early wake | 16 | 2.08 | Insomnia | Dichotomized |
| ESS | 34 | 4.43 | Insomnia | Continuous |
| ESS>10 (EDS) | 8 | 1.04 | Insomnia | Dichotomized |
| difficulty fall asleep | 17 | 2.21 | Insomnia | Dichotomized |
| frequent wake | 0 | 0.00 | Insomnia | Dichotomized |
| taking sleep pill | 73 | 9.51 | Insomnia | Dichotomized |
| restless sleep | 6 | 0.78 | Insomnia | Dichotomized |
| WHIIRS | 35 | 4.56 | Insomnia | Continuous |
| avg event Duration | 0 | 0.00 | SDB | Continuous |
| hypoxic burden | 7 | 0.91 | SDB | Continuous |
| per90 | 1 | 0.13 | SDB | Dichotomized |
| rei0 | 11 | 1.43 | SDB | Continuous |
| rei0>15 | 4 | 0.52 | SDB | Dichotomized |
| rei0>5 | 27 | 3.52 | SDB | Dichotomized |
| total event count | 12 | 1.56 | SDB | Continuous |
| total event Duration | 7 | 0.91 | SDB | Continuous |
| rei3 | 68 | 8.85 | SDB | Continuous |
| rei3>15 | 8 | 1.04 | SDB | Dichotomized |
| rei3>5 | 43 | 5.60 | SDB | Dichotomized |
| min o2 | 38 | 4.95 | SDB | Continuous |
| avg o2 | 36 | 4.69 | SDB | Continuous |
| perlt90 | 8 | 1.04 | SDB | Continuous |
| snore | 4 | 0.52 | SDB | Dichotomized |
| weekday bed time | 158 | 20.57 | Timing | Continuous |
| weekday midpoint time | 197 | 25.65 | Timing | Continuous |
| weekday wake time | 153 | 19.92 | Timing | Continuous |
| **weekend bed time** | 80 | 10.42 | Timing | Continuous |
| **weekend midpoint time** | 81 | 10.55 | Timing | Continuous |
| **weekend wake time** | 64 | 8.33 | Timing | Continuous |

Among the 768 included metabolites, carbohydrates had the highest average number of statistically significant associations with sleep phenotypes (mean: 4.23, range: 1 – 8 per metabolite), followed by cofactors and vitamins (mean: 3.92, range 0 – 12 per metabolite) and lipids (mean: 3.04, range: 0 – 11 per metabolite). Partially characterized molecules (mean: 1.78, range: 0 – 8) and xenobiotics (mean: 1.78, range: 0 – 9) had the lowest number of statistically significant associations with sleep phenotypes per metabolite (Supplementary Table S3).

We aggregated metabolites by subpathway and sleep phenotypes by domain, then identified and visualized top metabolomic subpathways with over 25% statistically significant associations (defined as FDR-adjusted *p*<0.05) among all tested associations (Figure 4) by sleep phenotype domain. Primary bile acid metabolism showed the highest cumulative percentage of statistically significant associations across all five sleep phenotype domains, although no significant association was identified for SDB phenotypes (Supplementary Table S4). Both lipids and cofactors/vitamins had subpathways (e.g., ketone bodies, acyl glutamine, pantothenate and CoA metabolism, nicotinate and nicotinamide metabolism) that had high percentage of statistically significant associations with sleep timing phenotypes. SDB phenotypes had the highest percentage of statistically significant associations among lipids, especially progestin steroids, phosphatidylethanolamine, diacylglycerols, pregnenolone steroids and sphingomyelins, which is consistent with our previous research findings (43). As for the sleep duration domain, the top subpathways with the highest percentage of significant associations were bacterial/fungal, amino sugar metabolism, nicotinate and nicotinamide metabolism, primary bile acid metabolism, corticosteroids, and polyamine metabolism. Insomnia domain generally had fewer statistically significant associations, among which pregnenolone steroids and vitamin A metabolism showed the highest percentage of statistically significant associations.

**Figure 4.**
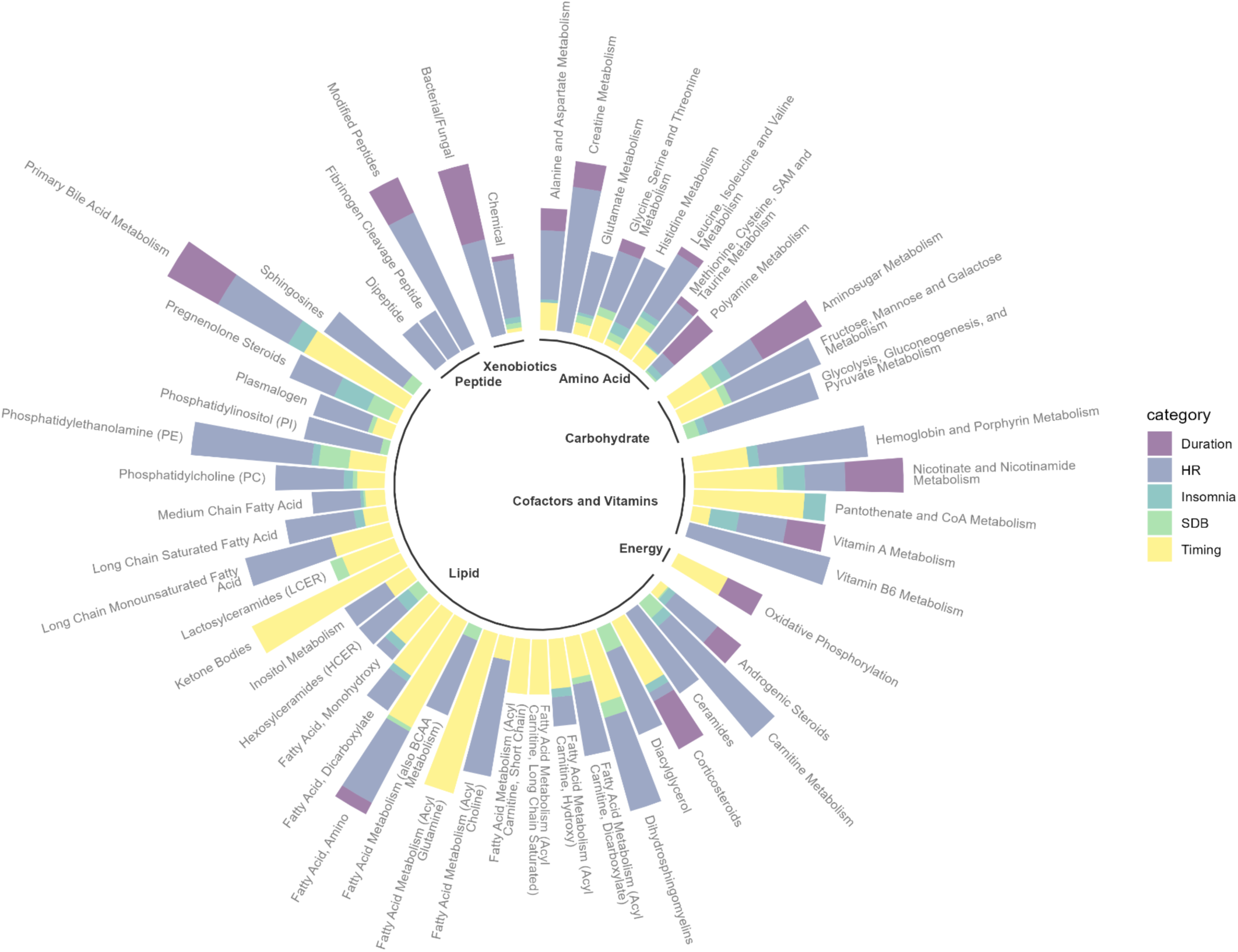
Number of statistically significant associations between metabolites and sleep phenotypes aggregated by subpathway and sleep phenotype domain.

Comparing men and women, potential sex differences can be observed (Supplementary Figure S2), with the limitation that the sample size of male participants was lower. Relatively more subpathways had statistically significant associations with SDB phenotypes among females compared to males (these include oxidative phosphorylation, lactosylceramides fructose, mannose and galactose metabolism). The subpathways from which metabolites were associated with insomnia phenotypes were mostly pregnenolone steroids, nicotinate and nicotinamide metabolism and carnitine metabolism among females, while long chain saturated and monounsaturated fatty acid and amino sugar metabolism were more associated with insomnia phenotypes among males.

We also aggregated metabolites by superpathway and visualized the percentage of statistically significant associations between superpathways and sleep phenotype domains in the format of a heatmap (Figure 5). The percentage of statistically significant associations were highest for HR during sleep, especially among carbohydrates (40.34%), followed by cofactors and vitamins (26.92%), amino acids (24.70%) and peptides (24.04%) (Supplementary Table S5). Insomnia and SDB domain showed overall lower percentage of statistically significant associations with metabolites across all superpathways, all of which were below 10%.

**Figure 5.**
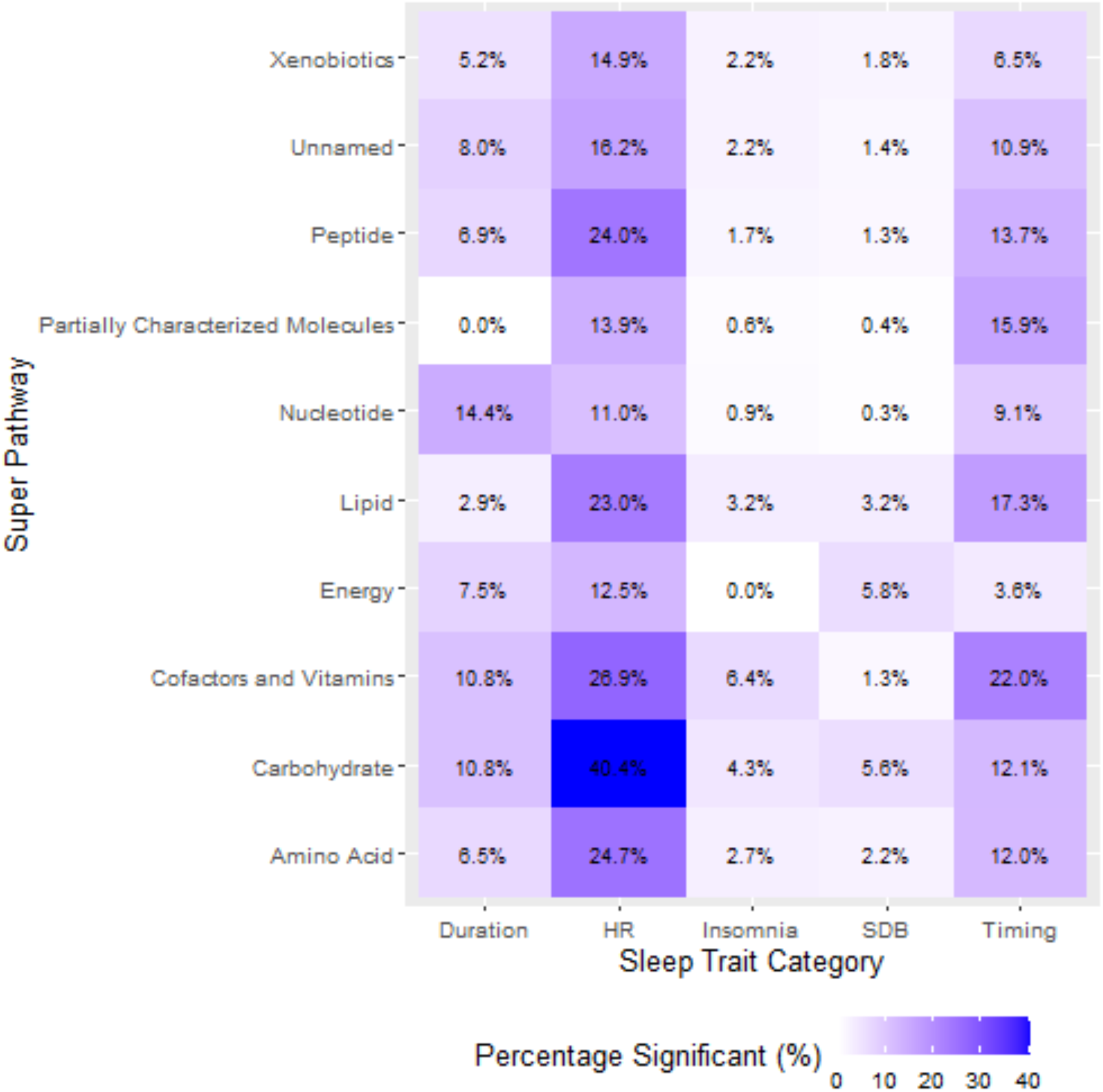
Percentage of statistically significant associations between metabolites and sleep phenotypes aggregated by superpathway and sleep phenotype domain.

Among all metabolites associated with at least one sleep phenotype, we identified the top 10% (n=37) with the highest number of total associations, regardless of domains (Figure 6). Sixteen metabolites were lipids, 8 were amino acids, 4 were cofactors and vitamins, 4 were unnamed, 2 were peptides, 1 was carbohydrate, 1 was xenobiotic and 1 was partially characterized. Every metabolite from the top 10% list had associations with more than one sleep phenotype domain, among which 11 metabolites were associated with four different domains of sleep phenotypes. The amino acids Vanillylmandelate (VMA) and 1-carboxyethylisoleucine were associated with the greatest number of sleep phenotypes from all sleep phenotype domains except for the insomnia domain.

**Figure 6.**
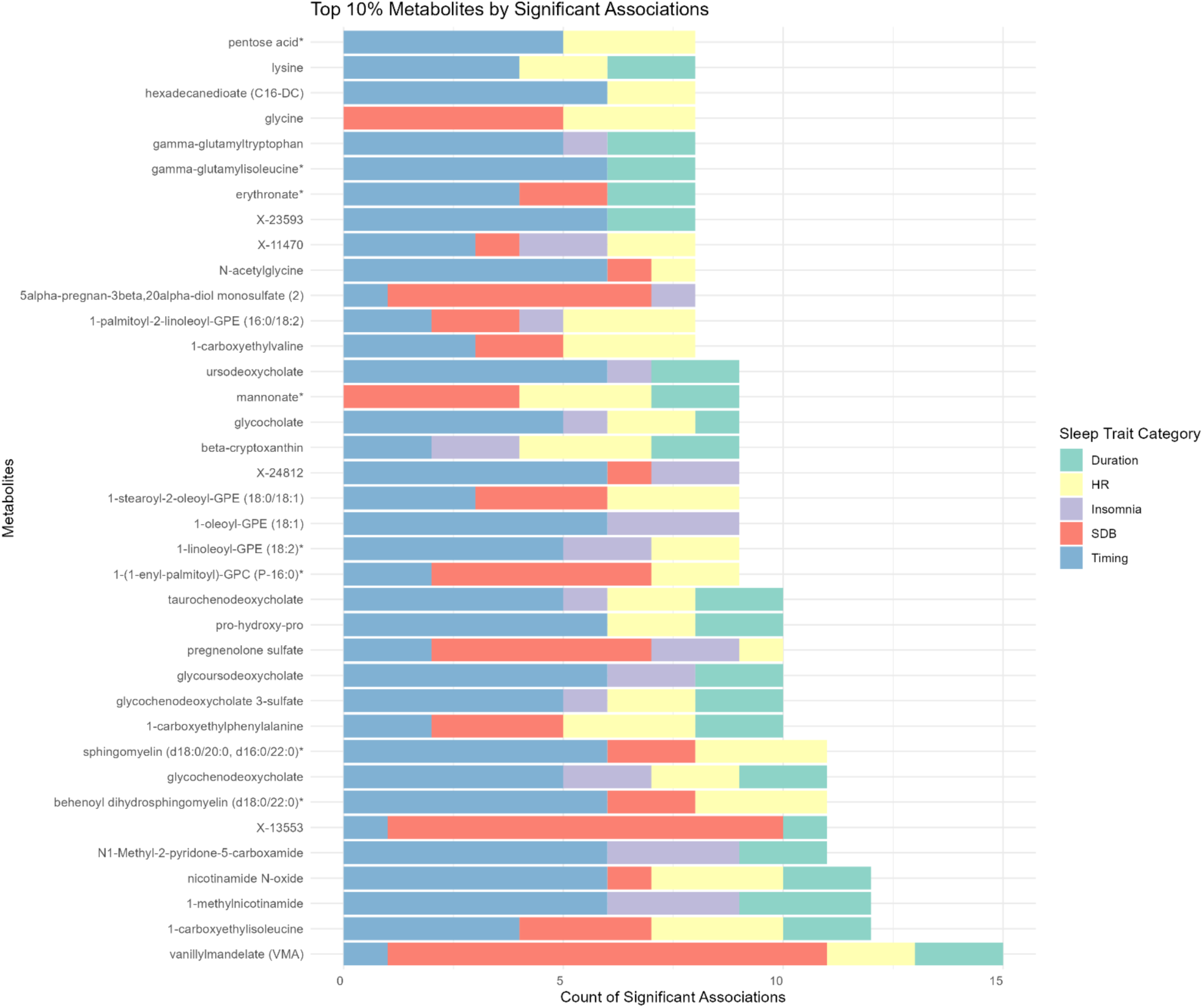
Number of significant associations aggregated by sleep phenotype domain among the top 10% connected metabolites with the most statistically significant associations.

We also compared the similarities among sleep phenotype domains by calculating the Dice Similarity Coefficients (DSC) which quantifies the associated metabolites overlap between any two sleep domains. As shown in Figure 7, the highest overlap was observed between HR during sleep phenotypes and SDB (DSC=0.35), and with sleep timing domain (DSC=0.35), respectively. The overlap between SDB and sleep timing domain was lower (DSC=0.26). High overlap was also observed between sleep duration and sleep timing (DSC=0.31). The lowest levels of overlap were observed for sleep duration traits and SDB domain (DSC=0.13) and sleep duration and HR during sleep phenotypes (DSC=0.19).

**Figure 7.**
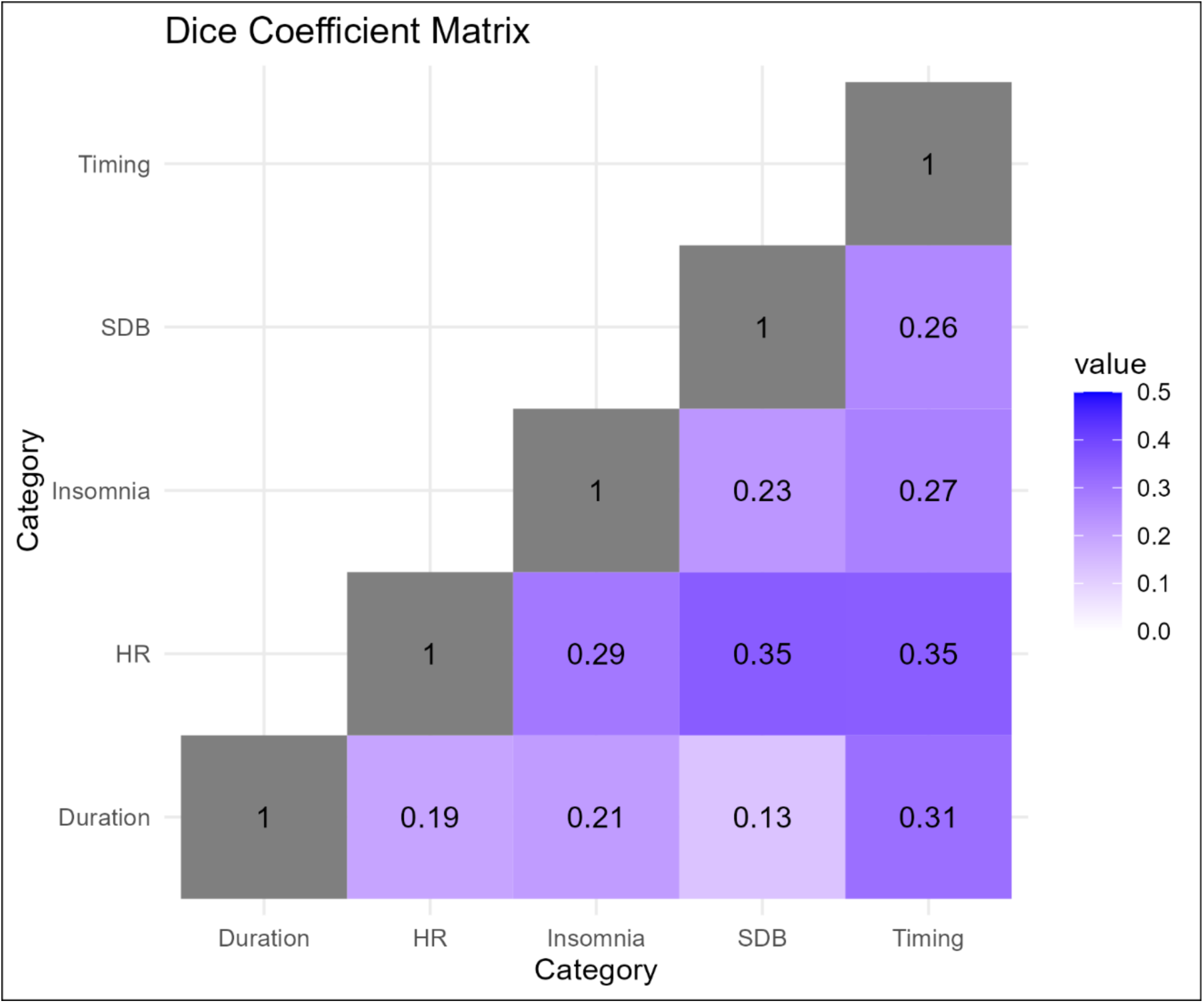
Dice coefficient matrix of sleep phenotype domain based on associated metabolites. Dice coefficient was calculated based on the shared associations with metabolites between any two sleep phenotype domain over the total significant associations for both sleep phenotype domains combined. A high value (maximum value of 1) indicates complete overlap, while a low value (minimum value of 0) indicates no overlap between two sleep phenotype domains.

## Bipartite network analysis based on statistically significant associations between metabolites and sleep phenotypes

Figure 8 shows a network of nodes and edges in which each node represents either a sleep phenotype domain or a metabolomic subpathway, while each edge represents one or more statistically significant associations between two nodes based on the single metabolite association analysis results (FDR-adjusted *p*<0.05). The width of the edge indicates the number of statistically significant associations between two nodes. Some metabolomic subpathways have exclusive connections with one sleep domain, such as fatty acid and dicarboxylate subpathways and sleep timing domain. Others tend to have many connections with multiple sleep domains, such as sphingomyelins with sleep timing, heart rate and SDB phenotype domain. The placement of the nodes reflects their connectivity similarities – the closer two nodes are, the more similar their overall connectivity patterns are. SDB and heart rate phetnoypes, for instance, are closely placed together which also corresponds to the high DSC values between the two groups (Figure 7). Metabolomic subpathways located close to the center of the network indicates their connectivity with multiple sleep domains, while metabolomic subpathways located at the outskirt of the network indicates their connections with one or few closely located sleep phenotype domains. For instance, the cluster of fatty acids (i.e., fatty acid, dicarboxylate, fatty acid metabolism (acyl carnitine, hydroxy), fatty acid metabolism (acyl choline), long chain saturated fatty acid, and fatty acid, monohydroxy, appearing in the left quadrant of Figure 8) are mostly connected to sleep timing, heart rate, and insomnia sleep domains, while lipids such as lysophospholipid, secondary bile acid metabolism and androgenic steroids are connected to sleep duration and SDB phenotype domain.

**Figure 8.**
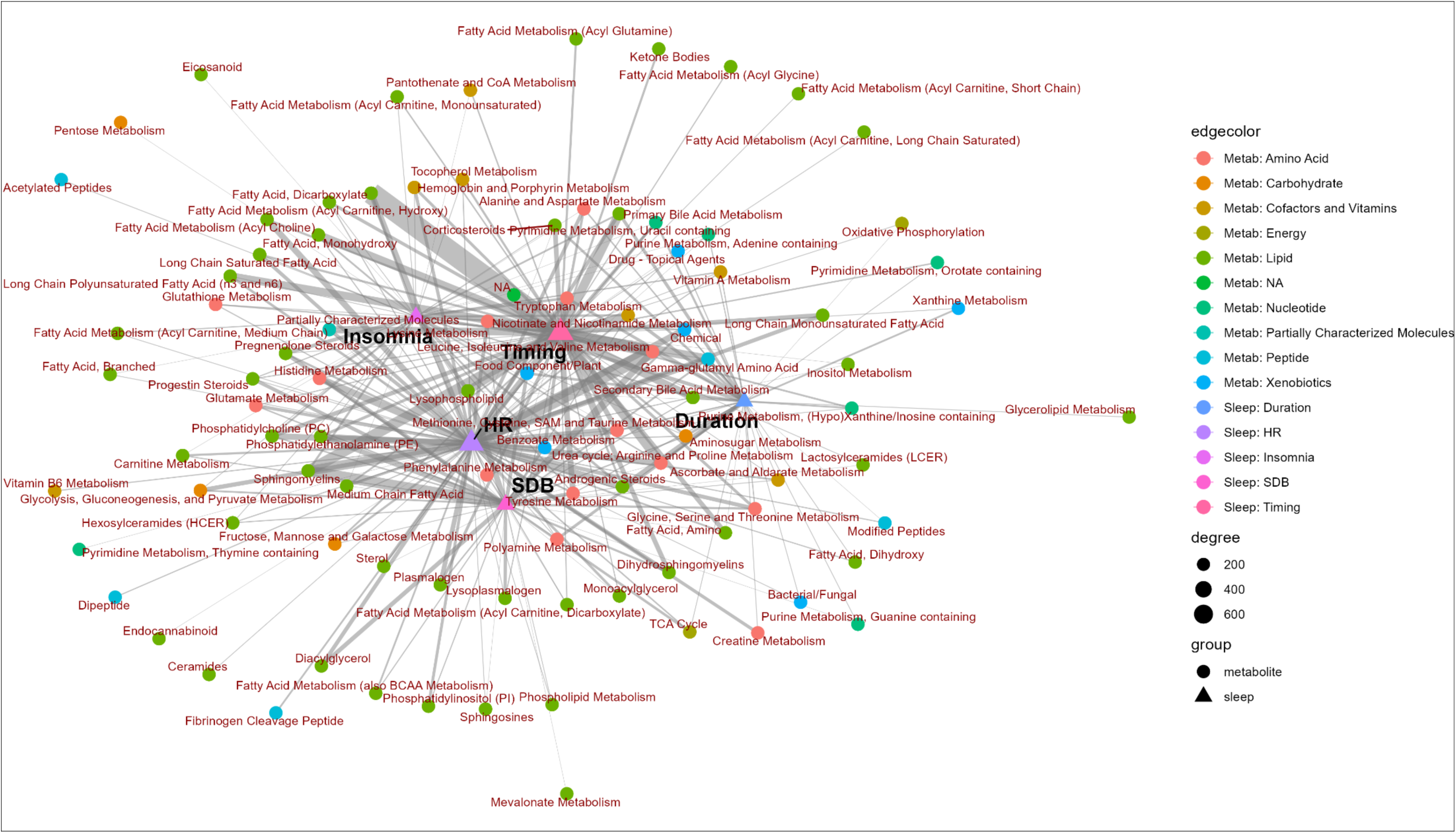
Network based on associations between metabolites and sleep phenotypes aggregated by subpathway and sleep phenotype domain using the Fruchterman-Reingold force-directed algorithm. Here we present a network of nodes and edges in which each node represents either a sleep phenotype domain or a metabolomic subpathway, while each edge represents one or more statistically significant associations between two nodes based on the single metabolite association analysis results (FDR-corrected p<0.05). The width of the edge (i.e., degree) indicates the number of statistically significant associations between two nodes. The placement of nodes is based on connectivity similarity between nodes, where similar connectivity is reflected as proximity of nodes.

Treated as a bipartite network model, we further characterized the connectivity between the metabolites, grouped at the superpathway level, and sleep domains quantified by network properties. The number of connected metabolites is the lowest for energy metabolites and the highest for lipids, which corresponds to the number of connected sleep phenotypes for the two superpathways and the average connection per node (i.e., links per node) (Figure 9). Similar trend can be observed for the mean number of shared metabolites among sleep phenotypes – the number of overlapping metabolites for sleep phenotypes is the highest among lipids and lowest among energy metabolites, which suggests that more lipids are connected to the same sleep phenotype compared to metabolites from other superpathways. The mean number of shared sleep phenotypes among metabolites, however, peaked for carbohydrates and cofactors and vitamins, followed by lipids, suggesting metabolites from the former two superpathways are more likely to connected to similar sleep phenotypes than other superpathways, despite their low number of total metabolites.

**Figure 9.**
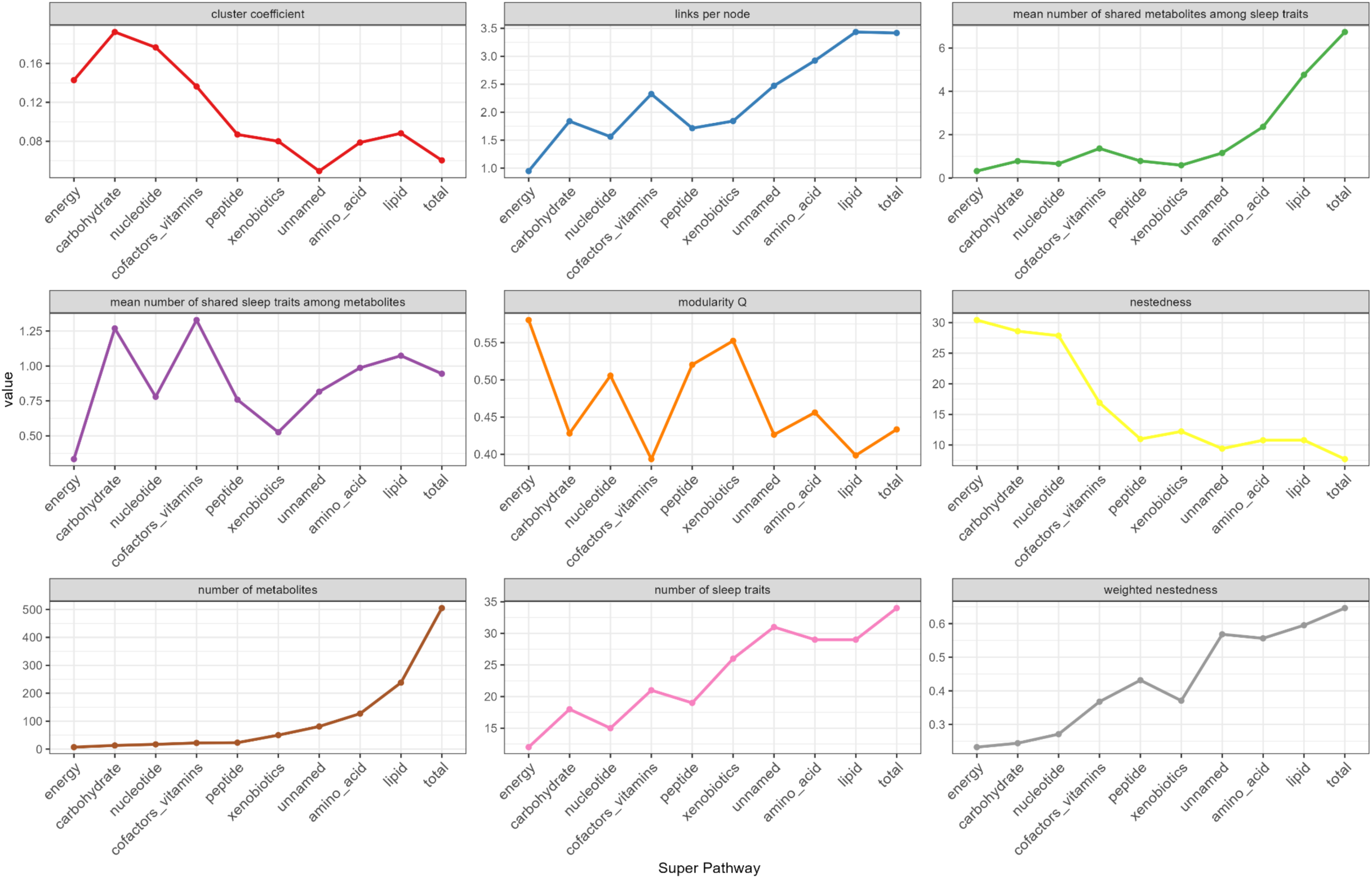
Network structural property metrics summarized by superpathway. Network properties metrics were calculated for the bipartite network based on the significant associations between metabolites and sleep phenotypes (FDR-corrected *P*<0.05). Cluster coefficients is the number of realized links divided by the number of possible links [https://www.rdocumentation.org/packages/bipartite/versions/2.19/topics/networklevel]. Nestedness measures how the interactions of less connected nodes are a subset of the interactions of more connected nodes. A value of 0 indicates high nestedness, while a value of 100 indicates “chaos”. Weighted nestedness considers interaction frequencies of the network, proposed by Galeano et al.(75). It ranges between 1 (perfect nestedness) and 0 (perfect chaos). Modularity Q is a measure to quantify how well a network can be partitioned into different groups of nodes such that nodes that belong to the same group are more likely to be connected than nodes that belong to different groups.

Cluster coefficient, the number of realized links divided by the number of possible links, is the highest among carbohydrates and nucleotides. This pattern indicates a higher degree of overlap in the neighboring nodes among carbohydrates and nucleotides, compared to unnamed metabolites and when combining all metabolites together regardless of superpathways, which may suggest a lower degree of diversity in terms of their connectivity patterns.

Nestedness of a graph is the property where groups of connected nodes are nested within larger groups of connected nodes. Two versions of nestedness measures were calculated. Both weighted (using the number of sleep domain-superpathway associations) and non-weighted nestedness measures generally agree, and suggest relatively high nestedness among lipids, amino acids, unnamed metabolites as well as combining all metabolites together, and relatively low nestedness among energy, carbohydrates and nucleotides (Supplementary Table S6).

Modularity Q is a measure of community structure. Modulatory values are higher when there are more clusters of connected nodes in a graph. Modularity Q was the highest among energy (Q=0.57) and xenobiotics (Q=0.57), followed by peptides (Q=0.52), and the lowest among cofactors and vitamins (Q=0.39) and lipids (Q=0.4), suggesting that among the former three superpathways, there are groups of metabolites in the superpathway that tend to connect to the same sleep phenotype. Among lipids, and cofactors and vitamins, there are less such “communities” of metabolites with similar sleep domain connections.

## Discussion

In this study we performed single metabolite association analyses for a variety of sleep phenotypes including SDB, insomnia, sleep duration, sleep timing, and heart rate measures during sleep, adjusted for common demographic covariates and BMI, and identified metabolites with statistically significant associations with each sleep phenotype. Taking a network analysis approach, we developed a bipartite network based on the identified associations between metabolites and sleep phenotypes. This approach characterizes the connectivity patterns of metabolites from different metabolite super- and sub-pathways, and identifies “neighboring” metabolites and sleep phenotypes that are inter-connected, as well as metabolomic subpathways that may be of interest for further study.

Many of the metabolites identified in our study as associated with sleep phenotypes have been previously reported to be linked to various sleep phenotypes, usually concurring with our findings. Among the top 10% of connected metabolites with the highest number of associations, regardless of sleep phenotype domain, almost all metabolites were reported in prior publications (Table 3), of which 22 were related to sleep. These include some published results from our prior work in HCHS/SOL (i.e., non-independent findings). Glycine, for instance, has shown associations with multiple SDB phenotypes, and was reported to be associated with sleep deprivation (44,45). In a recent study, glycine was found to be depleted in blood among Hispanic/Latino individuals with severe SDB, likely due to microbiome changes in an oxygen-poor environment (46). Four other metabolites – 1-oleoyl-GPE (18:1), 1-(1-enyl-palmitoyl)-GPC (P-16:0)*, pregnenolone sulfate, 5alpha-pregnan-3beta,20alpha-diol monosulfate (2), previously reported to be associated with novel SDB phenotype metrics after dimension reduction in our prior work in HCHS/SOL (43), were here also associated with several SDB phenotypes as well as with phenotypes from domains including insomnia, sleep timing, and heart rate during sleep.

**Table 3:** Top sleep-connected metabolites and their relevant previously-reported associations.

| **Metabolite** | **Super pathway** | **Sub pathway** | **Prior publications** |
| --- | --- | --- | --- |
| **glycine** | Amino Acid | Glycine, Serine and Threonine Metabolism | Sleep deprivation (44), OSA (46) |
| **N-acetylglycine** | Amino Acid | Glycine, Serine and Threonine Metabolism | Sleep restriction (53) |
| **1-carboxyethylisoleucine** | Amino Acid | Leucine, Isoleucine and Valine Metabolism | T2DM(47) and GDM (48) |
| **1-carboxyethylvaline** | Amino Acid | Leucine, Isoleucine and Valine Metabolism | HTN (49) |
| **lysine** | Amino Acid | Lysine Metabolism | Insomnia (54), repeated sleep disruption (55) |
| **1-carboxyethylphenylalanine** | Amino Acid | Phenylalanine Metabolism | T2DM (47) |
| **vanillylmandelate (VMA)** | Amino Acid | Tyrosine Metabolism | Sleep deprivation (56) |
| **pro-hydroxy-pro** | Amino Acid | Urea cycle; Arginine and Proline Metabolism | Sleep midpoint (27) |
| **erythronate\*** | Carbohydrate | Aminosugar Metabolism | Sleep midpoint and wake time (57) |
| **1-methylnicotinamide** | Cofactors and Vitamins | Nicotinate and Nicotinamide Metabolism | Sleep restriction (58) |
| **N1-Methyl-2-pyridone-5-carboxamide** | Cofactors and Vitamins | Nicotinate and Nicotinamide Metabolism | Sleep restriction (58) |
| **nicotinamide N-oxide** | Cofactors and Vitamins | Nicotinate and Nicotinamide Metabolism | Anti-inflammatory effect (59) |
| **beta-cryptoxanthin** | Cofactors and Vitamins | Vitamin A Metabolism | Sleep duration (60), sleepiness and sleep disturbance (61) cognitive function (52) |
| **behenoyl dihydrosphingomyelin (d18:0/22:0)\*** | Lipid | Dihydrosphingomyelins | Physical exercise (62) |
| **sphingomyelin (d18:0/20:0, d16:0/22:0)\*** | Lipid | Dihydrosphingomyelins | Blood pressure (63) |
| **hexadecanedioate (C16-DC)** | Lipid | Fatty Acid, Dicarboxylate | Blood pressure regulation (50), neurodegenerative diseases (51) |
| **1-linoleoyl-GPE (18:2)\*** | Lipid | Lysophospholipid | Depression (64) |
| **1-oleoyl-GPE (18:1)** | Lipid | Lysophospholipid | SDB (43) |
| **1-(1-enyl-palmitoyl)-GPC (P-16:0)\*** | Lipid | Lysoplasmalogen | SDB (43) |
| **1-palmitoyl-2-linoleoyl-GPE (16:0/18:2)** | Lipid | Phosphatidylethanolamine (PE) | Infant sepsis (65) |
| **1-stearoyl-2-oleoyl-GPE (18:0/18:1)** | Lipid | Phosphatidylethanolamine (PE) | OSA (24) |
| **pregnenolone sulfate** | Lipid | Pregnenolone Steroids | SDB (43), REM sleep (66) |
| **glycochenodeoxycholate** | Lipid | Primary Bile Acid Metabolism | Poor sleep quality (67) |
| **glycochenodeoxycholate 3-sulfate** | Lipid | Primary Bile Acid Metabolism | OSA (29) |
| **glycocholate** | Lipid | Primary Bile Acid Metabolism | OSA (26) |
| **taurochenodeoxycholate** | Lipid | Primary Bile Acid Metabolism | OSA (26), sleep midpoint (27) |
| **5alpha-pregnan-3beta,20alpha-diol monosulfate (2)** | Lipid | Progestin Steroids | SDB (43) |
| **glycoursodeoxycholate** | Lipid | Secondary Bile Acid Metabolism | Bed time (68) |
| **ursodeoxycholate** | Lipid | Secondary Bile Acid Metabolism | OSA intermittent hypoxia/hypercapnia (69) |
| **pentose acid\*** | Partially Characterized Molecules | Partially Characterized Molecules | REM vs wakefulness (70) |
| **gamma-glutamylisoleucine\*** | Peptide | Gamma-glutamyl Amino Acid | Sleep midpoint (27), wake time and sleep midpoint (57) |
| **gamma-glutamyltryptophan** | Peptide | Gamma-glutamyl Amino Acid | Late-onset sepsis among premature infants (71) |
| **mannonate\*** | Xenobiotics | Food Component/Plant | Undernutrition (72) |

Some of the metabolites identified in this study were also reported to be associated with other comorbidities such as cardiovascular, metabolic, and neurodegenerative diseases, potentially connecting sleep to a wide range of chronic adverse health outcomes. For instance, 1-carboxyethylisoleucine and 1-carboxyethylphenylalanine, both associated with sleep duration, SDB, sleep timing and heart rate during sleep, were both reported to be altered in blood among type II diabetes patients (47), while 1-carboxyethylisoleucine, among several other branched-chain amino acid metabolites, was also altered in serum levels among gestational diabetes patients (48). Another amino acid from the top 10% connected metabolites list, 1-carboxyethylvaline, was reported to be associated with hypertension (49). Hexadecanedioate (C16-DC), a dicarboxylate fatty acid, was reported to be associated with blood pressure regulation (50) and neurodegenerative diseases (51). Beta-cryptoxanthin, an antioxidant and pre-vitamin A carotenoid found in fruits and vegetables, was found positively associated with cognition (analysis was not adjusted for sleep traits) in individuals of diverse race/ethnic backgrounds (52). These metabolites suggest some shared biochemical mechanisms between sleep and other chronic adverse health outcomes.

When working with complex data such as untargeted metabolomic profiling and multiple phenotypes, effectively summarize data and develop useful insights is challenging (73). Different visualization and analytical approaches have been developed to facilitate this process (74). Here, we applied a systems biological approach – network analysis which has been widely used in gene expression, gene regulation, gene-disease network, and drug-drug interaction studies, on statistical relations among multiple phenotypes and metabolites. This data-driven network approach is different from knowledge-based network construction approaches (75) built on biochemical relations such as KEGG networks (76). Here, the network was built on inferred statistical relations among metabolites and phenotypes, leveraging, for interpretation purposes, well-studied network properties metrics from other fields such as ecology, socioeconomics, neuroscience, drug-disease networks, among others (77–80). One interesting observation is the relatively high nestedness among lipids and amino acids. High nestedness indicates that metabolites from these two superpathways form such a structure that metabolites with fewer connections (referred to as “degree”) are more likely to “connect”, via their mutual connections with sleep domains, with metabolites with higher degree of connections, rather than with other metabolites with a similar low degree of connectiveness. An intuitive depiction of such network is a “core-periphery” structure, in which a “core” of nodes is connected with other nodes, where a “periphery” of nodes tends to only connect with the nodes in the core. Contextually, among lipids and amino acids associated with sleep phenotypes, higher nestedness, compared to other superpathways, implies that these superpathways are more likely to have a subset of metabolites that play a “key” role forming connections with many, and the same, sleep phenotypes, rather than an evenly distributed network where metabolites form connections with various sleep phenotypes from different domains in a random manner. This is supported by the pattern observed in Figure 8 where subpathways from amino acids and lipids superpathways tend to be placed in the center of the network graph. Metabolites with high connectiveness from these two super-pathways likely have roles in shared biological processes across multiple sleep phenotypes, especially considering 24 out of 37 top 10% connected metabolites are either amino acids or lipids. Modularity metrics, on the other hand, measure how well a network can be partitioned into clusters or compartments in which there are dense connections internally and sparser connections with other clusters (81). Among energy and peptide metabolites associated with sleep phenotypes, relatively higher modularity compared to other superpathways suggests the existence of such subgroups with distinct relationships with sleep phenotypes.

HR domain had the highest percentage of metabolite associations of these assessed. HR during sleep reflects activity of the autonomic nervous system, and is influenced by cardiac function, sleep stage (i.e., lowest in deep restorative sleep and highest and most variable in REM sleep and wakefulness during the sleep period), and sleep apnea-related heart rate response (82,83). Notably, both low and elevated heart rate response to SDB events are associated with biomarkers of cardiovascular disease, while elevated heart rate response to SDB events is predictive of incident fatal and non-fatal CVD (84). The finding that HR during sleep associated with hundreds of metabolites, from all superpathways, may reflect the multiple biological mechanisms that underlie this phenotype, as well as support the sleep-related HR as a marker of multiple biologically processes that may be targeted for interventions. Sleep timing also was associated with a relatively high number of metabolite associations, supportive of growing data implicating timing-related behaviors on metabolic outcomes (85), as well as the correlation of sleep timing with other sleep domains (86). In contrast, fewer associations were observed between SDB phenotypes and metabolites. This may be because that SDB measured by traditional metrics such as the AHI may poorly characterize disease that is influenced by multiple mechanistic pathways (87).

An analytic choice that we made that is worth discussing is missing metabolite data imputation. Here, we imputed missing values of metabolites with no more than 25% missing values. For non-xenobiotic metabolites, we selected the imputation method based on an empirical investigation of the proportion of replicated associations between batches, and the selected method was multiple imputation that included all metabolite and other measures (covariates and lab values) that have strong associations with metabolite levels. It is natural to question whether this may somehow bias results: for example, is it possible that, for instance, using diabetes and BMI (among the rest) to impute metabolite values will lead to metabolite values that are “too reflective” or BMI and diabetes and therefore will somehow generate spurious metabolite associations with sleep measures that are associated with BMI and diabetes? The answer is that this is very unlikely. Generation of metabolite values that are overly similar to BMI or diabetes (in this example) by the predictive mean matching function suggests overfitting to the values of these covariates. Such overfitting will reduce the likelihood of replication of associations in the second batch, rather than increase it. Further, the predictive mean matching has some randomness due to sampling, further limiting overfitting to the variables used in imputation. Finally, it is important to note that the correlation between metabolites and covariates, as with sleep, is an inherent characteristic of this biological signal (i.e. it is a “feature, not a bug”), as are the metabolite associations with sleep measures. Therefore, leveraging this characteristic is useful.

There are a few strengths to this study: we looked at multiple sleep phenotypes simultaneously which provides a unique opportunity for pattern recognition across phenotypes. We also explored the network analysis approach in summarizing the metabolite-sleep phenotypes associations which enabled us to leverage well developed network property metrics from other fields to offer new insights and potentially lead to more hypothesis generation. Additionally, we compared the metabolite-sleep phenotype associations in combined sexes and sex-stratified study populations, recognizing more work is needed to further understand the implications of potential differences. Lastly, the atlas created in this study will be a useful resource for the scientific community. This study also has a few limitations. First, our study population is based on the HCHS/SOL cohort, representative of the Hispanic/Latino population in the US. Although it’s important to study under-represented populations such as Hispanic/Latino individuals, further studies on other populations are needed to increase the generalizability of the findings. Second, the network analysis conducted in this study is not to make network inference but to summarize the associations results. Third, due to the large breadth of analyses we did not account for medication use in this work. This can influence associations as medications can affect metabolite levels and sleep phenotypes. Similarly, fourth, our primary model for which we summarize the results was not adjusted for comorbidities. In all, one cannot infer mechanisms and directionality of associations from these analyses. Fifth, another limitation is that the objective overnight sleep measures did not use EEG, limiting SDB (e.g. we do not have measures of arousals) as well as potential sleep staging measures. Finally, any comparison between the sleep domains used in this analysis is limited by the available sleep phenotypes, their number, and the correlation patterns between them.

In summary, we studied the associations between multiple sleep phenotypes from multiple sleep domains and the metabolomic environment in a large population-based cohort study. Using network analysis, we were able to visualize the interconnectedness between multiple sleep phenotypes and associated metabolites simultaneously, which provides an opportunity to glean into connectivity patterns that otherwise might be obscure when presented as individual relationships. We also created a resource for the sleep research community that will facilitate hypothesis generation in future metabolomic studies on sleep health. As sleep is highly affected by the social and built environment, it would be important, in the future, to use metabolomics to glean into the pathways by which the environment impacts sleep.

**Supplementary Figure S1: Study sample selection and pre-processing for the metabolomic analysis.**

**Supplementary Figure S2: Number of statistically significant associations between metabolites and sleep phenotypes aggregated by subpathway and sleep phenotype domain stratified by sex.**

## Data availability

HCHS/SOL data are available through application to the data base of genotypes and phenotypes (dbGaP) accession phs000810. HCHS/SOL metabolomics data are available via data use agreement with the HCHS/SOL Data Coordinating Center (DCC) at the University of North Carolina at Chapel Hill, see collaborators website: https://sites.cscc.unc.edu/hchs/. The metabolite association data generated in this study are provided in a shiny app https://bws-bidmc.shinyapps.io/20240410_for_shinyappsio/.

## Code availability

The code used in this work has been deposited in the public repository https://github.com/yzhang104/HCHS_SOL_Sleep_Metabolomics_Atlas.git.

## Ethics statement

The HCHS/SOL was approved by the institutional review boards (IRBs) at each field center, where all participants gave written informed consent, and by the Non-Biomedical IRB at the University of North Carolina at Chapel Hill, to the HCHS/SOL Data Coordinating Center. All IRBs approving the HCHS/SOL study are: Non-Biomedical IRB at the University of North Carolina at Chapel Hill. Chapel Hill, NC; Einstein IRB at the Albert Einstein College of Medicine of Yeshiva University. Bronx, NY; IRB at Office for the Protection of Research Subjects (OPRS), University of Illinois at Chicago. Chicago, IL; Human Subject Research Office, University of Miami. Miami, FL; Institutional Review Board of San Diego State University, San Diego, CA. All methods and analyses of HCHS/ SOL participants’ materials and data were carried out in accordance with human subject research guidelines and regulations. This work was approved by the Mass General Brigham IRB and by the Beth Israel Deaconess Medical Center Committee on Clinical Investigations.

## Declaration of interests

Dr. Redline discloses consulting relationships with Eli Lilly Inc. Additionally, Dr. Redline serves as an unpaid member of the Apnimed Scientific Advisory Board, as an unpaid board member for the Alliance for Sleep Apnoea Partners, and has received loaned equipment for a multi-site study: oxygen concentrators from Philips Respironics and polysomnography equipment from Nox Medical.

## Acknowledgements

The authors thank the staff and participants of HCHS/SOL for their important contributions. Investigators website - http://www.cscc.unc.edu/hchs/. This work was supported by National Heart Lung and Blood Institute (NHLBI) grants R01HL161012 to TS, R35HL135818 to SR, and National Institute on Aging grant R01AG80598 to TS. Support for metabolomics data was graciously provided by the JLH Foundation (Houston, Texas) and by NHLBI grant R01HL141824. The Hispanic Community Health Study/Study of Latinos is a collaborative study supported by contracts from the National Heart, Lung, and Blood Institute (NHLBI) to the University of North Carolina (HHSN268201300001I / N01-HC-65233), University of Miami (HHSN268201300004I / N01-HC-65234), Albert Einstein College of Medicine (HHSN268201300002I / N01-HC-65235), University of Illinois at Chicago (HHSN268201300003I / N01-HC-65236 Northwestern Univ), and San Diego State University (HHSN268201300005I / N01-HC-65237). The following Institutes/Centers/Offices have contributed to the HCHS/SOL through a transfer of funds to the NHLBI: National Institute on Minority Health and Health Disparities, National Institute on Deafness and Other Communication Disorders, National Institute of Dental and Craniofacial Research, National Institute of Diabetes and Digestive and Kidney Diseases, National Institute of Neurological Disorders and Stroke, NIH Institution-Office of Dietary Supplements.

## Contributions of the authors

Ying Z performed association analyses, network analyses, and data visualization. BWS developed the R/shiny app for visualization of results from single metabolite association analysis. Yu Z performed data imputation. DAW provided code for modeling circular sleep phenotypes. BY, QQ, and EB designed and established the metabolomics ancillary studies. MA, LA-S, MD, RK, and JC participated in HCHS/SOL study design, recruitment, and operations. Ying Z and TS drafted the manuscript. BWS, Yu Z, DAW, BY, QQ, MA, LA-S, EB, MD, RK, JC, and SR critically reviewed and approved the manuscript. TS supervised the work.

