## Supplementary text and figures for "Untargeted Metabolome Atlas for Sleep Phenotypes in the Hispanic Community Health Study/Study of Latinos"

### Supplementary Note 1: Sleep phenotypes used.

We used sleep measures assigned into 5 domains: sleep duration, heart rate during sleep, insomnia, sleep disordered breathing, and sleep timing. We provide details about these phenotypes by domain.

**Sleep duration:** questionnaire items included bed time and wake time over the weekdays and weekend days. Based on these, we computed sleep duration during weekdays, sleep duration during weekdays, sleep duration (weighted average of weekdays and weekend sleep duration, with weights being 5/7 and 2/7 for weekdays and weekend days, respectively), and binary variables for short sleep ($\leq 5$ hours) during weekdays, and long sleep ($\geq 9$ hours) during weekdays.

**Heart rate (HR):** minimum, maximum, average, and standard deviation of heart rate during sleep (all continuous measures).

**Insomnia:** this category included variables measured via the women health initiative insomnia rating scale (WHIIRS), which combines 4 likert scale questions, use of sleeping pills, restless sleep, and excessive daytime sleepiness. Specifically, we included the WHIIRS score as a continuous measure, the Epworth sleepiness scale (ESS; as a continuous measure) and questions dichotomized to binary: typical night's sleep in past 4 weeks (1 if restless or very restless, 0 otherwise), trouble getting back to sleep (1 if 3 or more times a week), wake up earlier than you plan (1 if 3 or more times a week), wake up several times at night (1 if 3 or more times a week), trouble falling asleep (1 if 3 or more times a week), taking sleeping pills (1 if 3 or more times a week), and also excessive daytime sleepiness (EDS) which dichotomized ESS as ESS>10 defining EDS.

**Sleep disordered breathing (SDB):** Multiple measures were based on counts of respiratory events during sleep. Certified polysomnologists manually edited artifacts, identified periods of sleep, and annotated each respiratory event with its associated oxyhemoglobin desaturation. Respiratory events were identified as a 50% or greater reduction in airflow lasting greater than or equal to 10 seconds. The respiratory event index (REI) is the number of respiratory events per estimated sleep hour, with the REI0 (“all desat”) and REI3 (“3% desat”), comprised of hypopneas scored without a desaturation requirement (REI0) or with associated desaturations of greater than or equal to 3% (REI3). We also used measures based on oxyhemoglobin saturation (SpO2) during sleep, including related to respiratory events. Specifically:

Continuous measures included REI0, REI3, total count of respiratory events, total length of respiratory events (i.e. the total sleep time with an event), average respiratory event length, hypoxic burden, minimum and average SpO2, percent sleep time with SpO2<90% (perlt90), and dichotomized variables: REI0 and REI3 $\geq 5$ and $\geq 15$ (each applied with both thresholds), and also self-reported snoring (1 if 6-7 times a week, 0 otherwise).

**Sleep timing:** all these variables were treated as circular, and included weekday wake and bed time, weekend wake and bed time, weekday sleep midpoint, weekend sleep midpoint, and social jetlag, defined as the difference between the sleep midpoint on weekends and on weekdays.

### Supplementary Note 2: Assessment of metabolite imputation strategy.

We compared a few approaches for metabolite imputation and transformation. To decide which approach to use in the analysis, we applied the various approaches on batch 1 and batch 2 metabolite datasets separately, performed association analyses using batch 1, and attempted replication testing of selected metabolites in batch 2. We recorded the number of single metabolite association being replicated in the association models adjusted for age, gender, center, Hispanic background and BMI. From batch 1 discovery analysis we took metabolites with False Discovery Rate (FDR)-adjusted p-value<0.05 for replication analysis in batch 2. For this investigation, an association was considered replicated in batch 2 if it had p-value<0.05 (unadjusted) in batch 2 analysis. We used the following sleep traits for this analysis: sleep duration, SD of heart rate during sleep, REI3%, midpoint of weekday sleep, wake time of weekday and weekend sleep.

We compared imputation approaches only for non-xenobiotics. For xenobiotics, we applied imputation with half of the minimum values of the corresponding metabolite. Thus, we focused on non-xenobiotic for this analysis. We compared imputation approaches. For all metabolites, and post-imputation transformation (rank-normalization or no transformation). Compared imputation included:

1. Imputation using half the minimum value observed for the metabolite in the batch.
2. Multiple imputations of the metabolite dataset using a multivariate model with a fully conditional specification ("metaonly").
3. Multiple imputations of the metabolite and covariates dataset using a multivariate model with a fully conditional specification ("metacov").
4. Multiple imputations of the metabolite dataset using a multivariate model where each metabolite is imputed using only the top 10 most highly correlated metabolites ("metaonly_UnMet").
5. Multiple imputations of the metabolite and covariates dataset using a multivariate model where each metabolite is imputed using only the top 10 most highly correlated metabolites and/or covariates ("metacov_UnMet").

The code used for the "UnMet" methods was taken from https://github.com/tofaquih/imputation_of_untargeted_metabolites, based on methodology reported in Faquih et al. (1). All implementations of multiple imputations used 5 imputed datasets, where association analyses were performed in each imputed dataset and later combined. Results are provided in Supplementary Table S7). Multiple imputation using all metabolite values and covariates and fully conditional specification, followed by rank normalization, was picked as the optimal method and was used in the main analysis which combined the two batches after imputation and rank-normalization.

### Supplementary Figure S1: Study sample selection and pre-processing for the metabolomic analysis

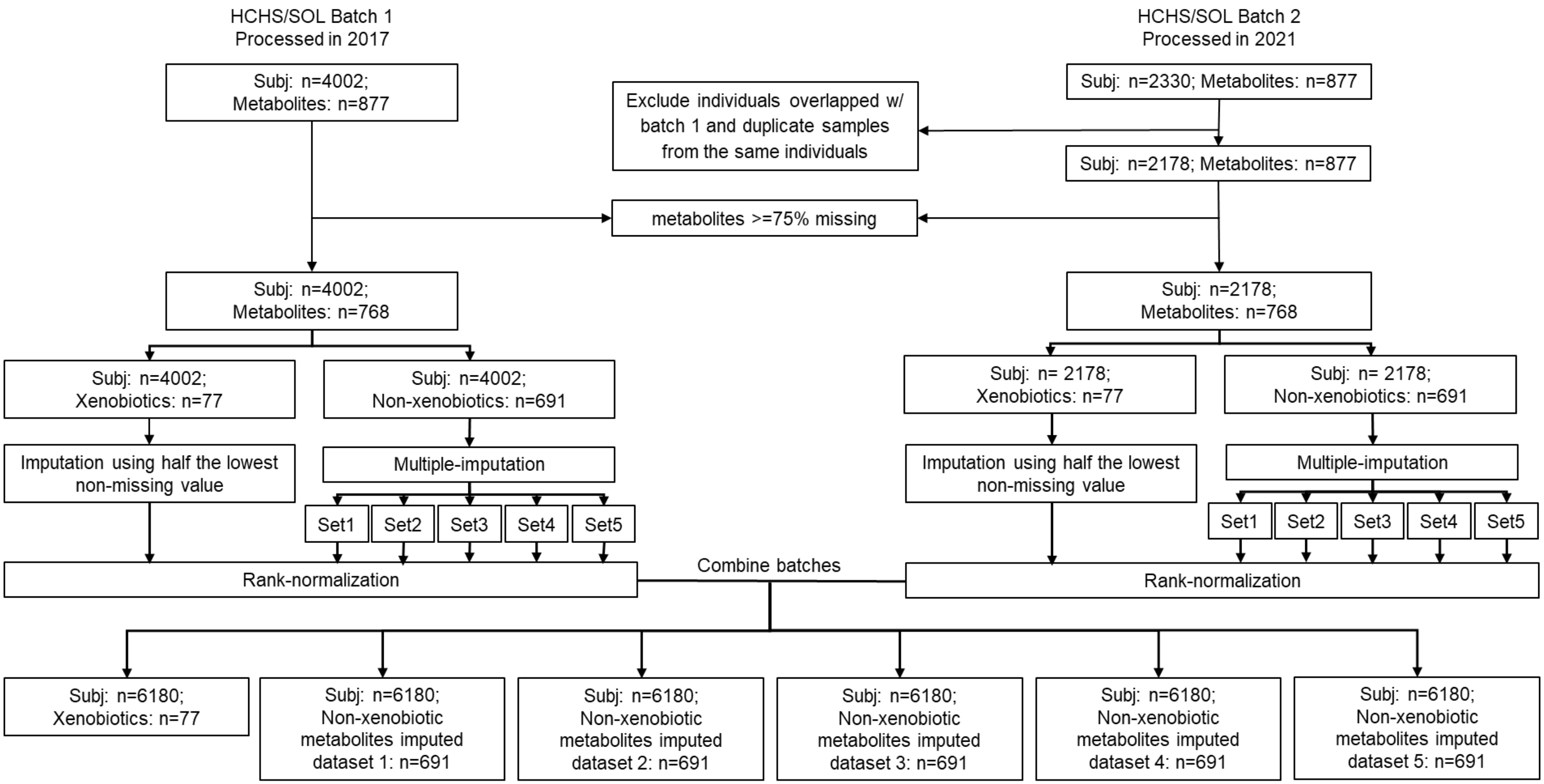

### Supplementary Figure S2: Number of statistically significant associations between metabolites and sleep phenotypes aggregated by subpathway and sleep phenotype domain stratified by sex

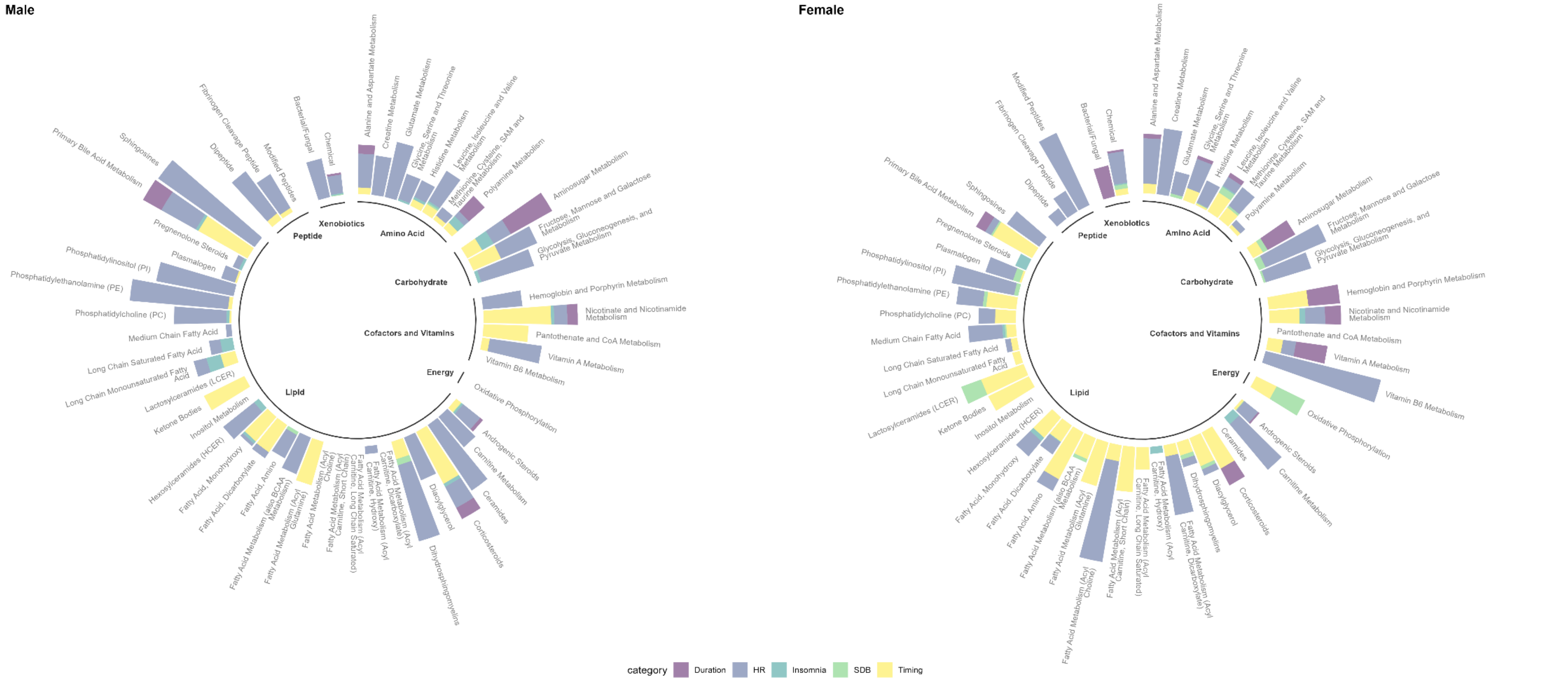
